# Longitudinal Measurement Properties of the Participation Questionnaire for Preschoolers: Reliability, Responsiveness, and Interpretability

**DOI:** 10.1101/2024.12.13.24318179

**Authors:** Takuto Nakamura, Iku Koshio, Sho Maruyama, Kohei Ikeda, Hirofumi Nagayama, Satoshi Sasada

## Abstract

**Purpose:** This study aimed to evaluate the test−retest reliability, responsiveness, and interpretability of the Preschool Questionnaire for Participation (PQP), a tool for assessing participation in preschool children with autism spectrum disorder (ASD).

**Methods:** A cohort study was conducted with data collected at four time points over six months from children with ASD aged 51–75 months. Test−retest reliability was examined using intraclass correlation coefficients (ICCs) for 275 participants who completed the study. Responsiveness was examined by analyzing correlations between changes in PQP scores and changes in multiple scales based on four predefined hypotheses. To assess interpretability, the Global Rating of Change (GRC) Scale was used as an anchor to analyze receiver operating characteristic curves and estimate the minimal important change (MIC).

**Results:** ICCs ranged from 0.80 to 0.92 for individual factors and reached 0.93 for the total score. Three of the four hypotheses for responsiveness were supported within the expected range. However, the correlation between the GRC Scale and changes in PQP scores was below 0.3, preventing MIC calculation.

**Conclusion:** While the MIC could not be calculated, the PQP demonstrated test−retest reliability equivalent to or better than existing participation measures. Given the limited verification of responsiveness for other tools, the PQP’s strong longitudinal reliability and validity provide a foundation for future research.

---

Participation is defined as the involvement in life situations and includes a wide range of everyday life involvement, such as “play,” “educational activities,” and “interpersonal relationships” (World Health Organization, 2001). Participation is an important intervention outcome for children with disabilities because it builds social connections and fosters independence. Children with autism spectrum disorder (ASD) who show limited social interaction and restricted interests have disability-specific restrictions in participation, such as difficulty participating in areas such as “educational activities” and “friendship” because of their disability characteristics (Yee et al., 2017). To accurately gauge the effectiveness of interventions for children with ASD, their participation must be measured in a disability-specific manner in both observational and intervention studies (Yee et al., 2017).

Several measurement tools have already been developed, and their reliability and validity have been verified to measure the participation of children with ASD. For example, The Structured Preschool Participation Observation for Children with ASD (SPO-ASD) is a measurement tool that allows occupational therapists to observe and evaluate the participation of children with ASD in educational situations, and it can be used in occupational therapy practice in schools (Golos et al., 2022). In addition, Picture My Participation - Chinese version (PMP-C) is a semi-structured interview using photographs. This child-centered measurement tool can clarify the areas of participation that require intervention based on the reports of children with ASD themselves (Li et al., 2023). Although it has not been verified for sufficient reliability and content validity, the Matrix for Assessment of Activities and Participation (MAAP) is the oldest tool that can comprehensively evaluate the participation of children with ASD through expert observation and interviews (Castro & Pinto, 2015).

As these measurement tools focusing on children with ASD have gradually developed, Nakamura, Nagayama, et al. (2024) developed the Parental Questionnaire for Participation (PQP), a participation measurement tool for caregivers to complete, intending to use it in large-scale surveys and as a simple tool in practice. The PQP, comprising 29 items in four factors, was developed through interviews with caregivers and experts in ASD (Nakamura, Koyama, et al., 2024), and scale development and cross-sectional measurement properties were verified through a questionnaire survey (Nakamura, Nagayama, et al., 2024). As a result of this research, acceptable internal consistency, structural validity, and construct validity of the PQP were reported.

However, the longitudinal measurement properties of the PQP have not been verified. The validation of the longitudinal measurement properties of a measurement tool is an essential process for understanding changes in scores over time. For example, the validation of one of the longitudinal measurement properties, test−retest reliability, and measurement error is an essential indicator for confirming the stability and consistency of scores when the same respondent answers at different points in time (Prinsen et al., 2018). In addition, the evaluation of responsiveness, which assesses the extent to which the tool is sensitive to changes in the underlying construct it measures, is an essential indicator for accurately grasping the effects of interventions and the efficacy of treatments (Prinsen et al., 2018). Further, interpretability is an indicator that enables us to understand the clinical or practical significance of the measurement results and supports appropriate judgment of the results and long-term decision-making (Prinsen et al., 2018).

Examining longitudinal properties in this way is essential for the PQP to better understand the participation of children with ASD and determine the extent to which exposure has changed their participation. Importantly, currently no studies have focused on longitudinal measurement properties specifically in children with ASD, highlighting a critical gap in understanding how their participation may change over time. Therefore, this study aims to examine the test−retest reliability, measurement error, responsiveness, and interpretability of the PQP.

## Methods

### Design

A prospective cohort study using an email survey was conducted from July 2023 to February 2024. Participants received a link to the online platform (Questant), where they provided informed consent. The study received approval from the XXX University of XXX Ethics Committee (Approval Number: XXX) and was preregistered prior to data analysis (XXX).

### Participants

Caregivers of children with ASD were asked to complete a questionnaire. The inclusion criteria were as follows: (1) the child has been diagnosed with ASD by a physician, (2) the child is a preschooler aged 51–75 months, and (3) the child has taken the Wechsler Intelligence Scale for Children, Wechsler Preschool and Primary Scale of Intelligence (Wahlstrom et al., 2018), Tanaka-Binet Intelligence Scale (Uno et al., 2014), or Kyoto Scale of Psychological Development (Koyama et al., 2009) and has an intelligence quotient (IQ) or developmental quotient (DQ) of 50 or higher. The exclusion criterion was as follows: the child has not been diagnosed with any disorder other than neurodevelopmental disorders.

### Measurement Tools

#### Parental Questionnaire for Participation

The PQP is a questionnaire designed for caregivers to measure the participation of children with ASD, with the following response options on a five-point Likert scale (Nakamura, Nagayama, et al., 2024): 5 (“Agree”), 4 (“Somewhat agree”), 3 (“Undecided”), 2 (“Somewhat disagree”), and 1 (“Disagree”). The PQP consists of 29 items regarding the child’s participation over the past three months. However, because this study was conducted concurrently with the scale development study, a trial version consisting of 36 items (including seven items later removed during item reduction analysis) was used. These seven deleted items were not reflected in the final scores. The PQP comprises four factors: “Factor 1: Friendship and Education,” “Factor 2: Family Satisfaction,” “Factor 3: Daily Life and Independence,” and “Factor 4: Leisure and Community Life.” The PQP has demonstrated acceptable reliability and validity. Scores for each factor are summed, with higher scores indicating better participation.

#### Strengths and Difficulties Questionnaire

The Strengths and Difficulties Questionnaire (SDQ) is a screening tool for children’s mental health (Matsuishi et al., 2008), comprising 25 questions addressing both positive and negative aspects of behavior. In this study, the Japanese parent version for children aged 4–16 years was used. Responses are rated on a three-point scale from “applies” to “does not apply.” The SDQ includes five factors: “Emotional Symptoms,” “Conduct Problems,” “Hyperactivity/Inattention,” “Peer Problems,” and “Prosocial Behavior.” The total difficulties score is calculated by summing the scores from all factors except “Prosocial Behavior.” Reliability and validity have been established, with higher scores on “Prosocial Behavior” indicating better outcomes, and higher scores on the other factors indicating worse outcomes.

#### Global Rating of Change Scale

The Global Rating of Change (GRC) Scale was used for two purposes: first, to confirm that the participant’s state of participation had not changed to verify test−retest reliability; second, to serve as an anchor for calculating the minimal important change (MIC) for the total PQP score and each factor score. The content of the GRC Scale needs to be tailored to the respondent’s needs (Kamper et al., 2009). In this study, the GRC Scale was designed to reflect the ICF definition of participation as “involvement in a life situation,” as shown in Table 1. Since participation is a multidimensional construct, multiple anchors were created to account for the possibility that a single-item question might not yield sufficient correlation coefficients with the PQP. Table 1 shows the GRC scales created, their purpose, and the time points at which they were used. The GRC Scale was administered only at the post-assessment time point, based on recommendations from the MIC study (Terwee et al., 2021). All GRC Scale responses were rated on a seven-point scale: 1, “Much worse;” 2, “Worse;” 3, “Slightly worse;” 4, “No change;” 5, “Slightly better;” 6, “Better;” and 7, “Much better.”

**Table 1.**
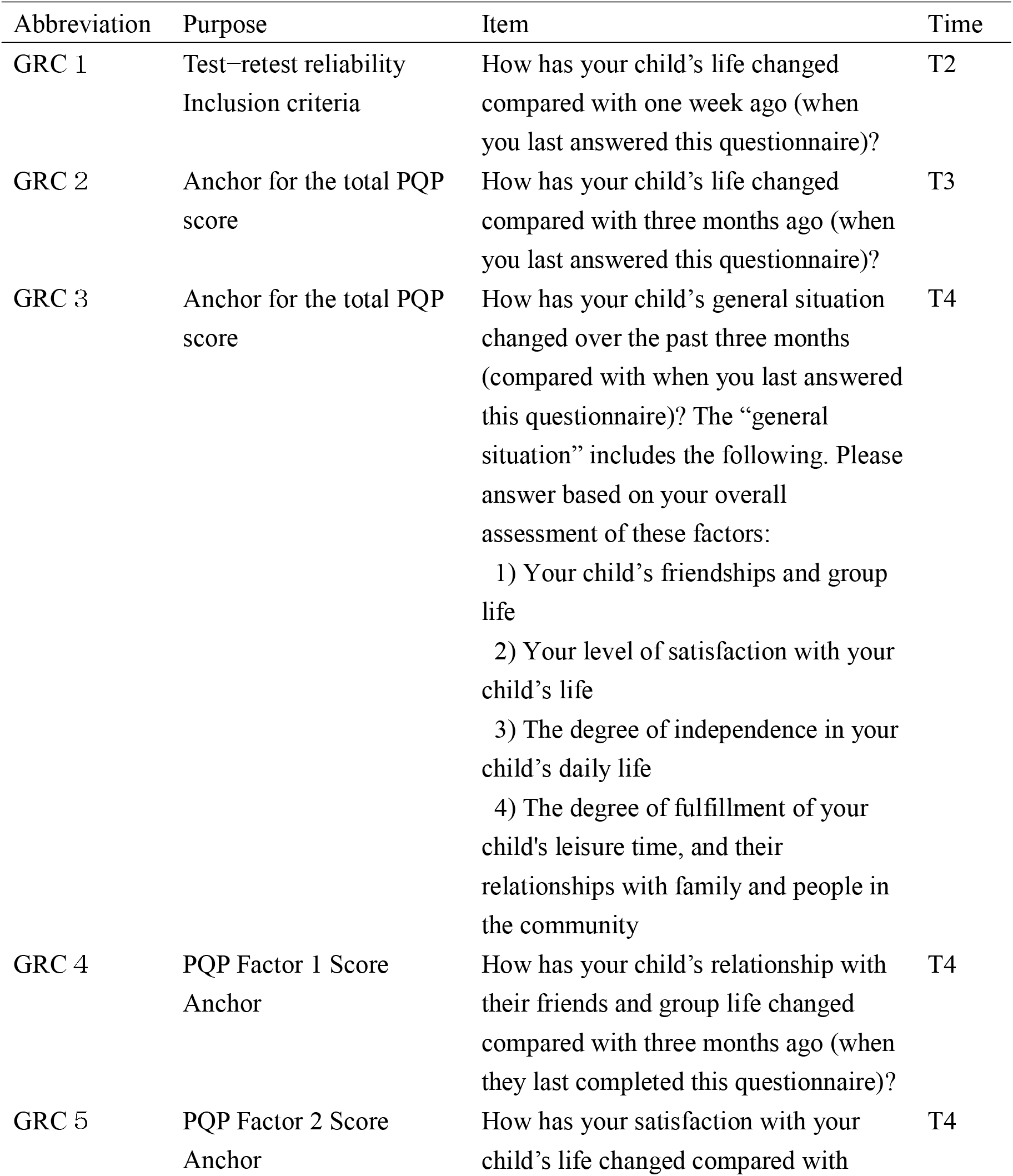
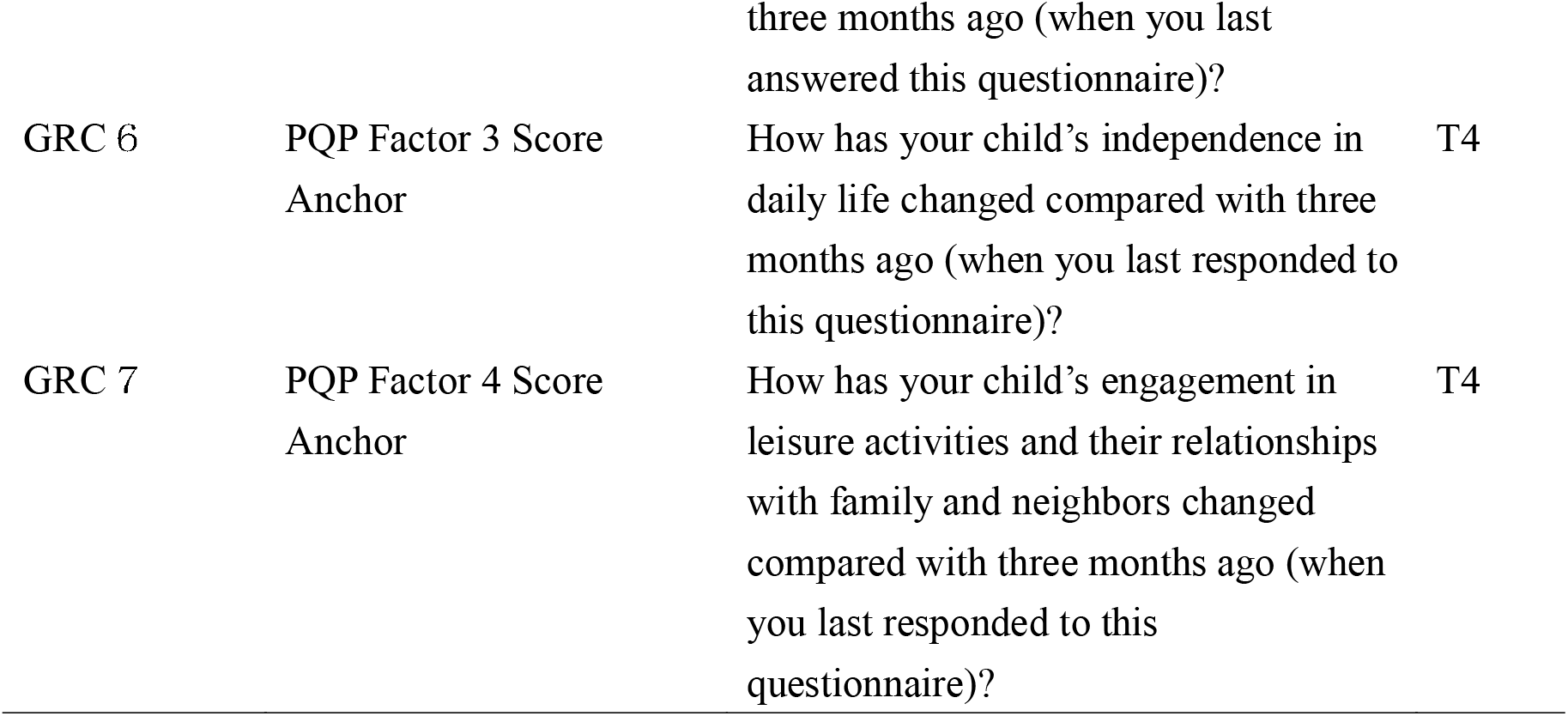
Details of Each Global Rating of Change Scale, Purpose of Use, and Data Collection Point in Time.

#### Social Responsiveness Scale, Second Edition

The Social Responsiveness Scale, Second Edition (SRS-2) is a questionnaire that assesses the severity of symptoms of ASD. It is designed to measure core symptoms of ASD in individuals aged from 2.5 years and older and comprises 65 questions (Constantino & Gruber, 2017). The child version (for ages 4–18 years) used in this study is answered by the caregiver, and the response options are on a four-point scale from “does not apply” to “almost always applies.” The total score is calculated from the five factors of “interpersonal awareness (SRS-Awr),” “social cognition (SRS-Cog),” “social communication (SRS-Com),” “social motivation (SRS-Mot),” and “restricted and repetitive behavior (SRS-RRB).” The total score is derived from the four factors other than “restricted and repetitive behavior (SRS-RRB).” The SRS-2 has been verified to have good reliability and validity. There are multiple cutoff points for the SRS-2, but the clinical cutoff points are 109.5 for boys and 102.5 for girls. A higher score indicates a higher severity of ASD.

#### Japanese Version of the Family Outcomes Survey-Revised

The Japanese Version of the Family Outcomes Survey-Revised (FOS-J) is the Japanese version of the FOS-R, a questionnaire that evaluates the results of support and intervention programs for children and their families (Ueda et al., 2015). This questionnaire consists of two sections: “Family Outcomes” and “Support Efficacy.” In this study, we used the “Family Outcomes” section, comprising five factors: “Understanding of child’s strengths, needs, and abilities,” “Understanding of own rights and advocacy for child,” “Assistance with child’s development and learning,” “Support system,” and “Access to community.” The response options range from “not at all true” to “always true” on a five-point scale, and a higher score indicates that the support benefits the family more effectively. Although the FOS-J has been verified for structural and construct validity, it has not yet been verified for reliability.

#### Demographic Questionnaire

The respondents were asked about their relationship to the child with ASD, their educational background, family structure, annual household income, place of residence, and their child’s gender, age, comorbid diagnoses, age at diagnosis, use of kindergarten/nursery school, use of medical and welfare services, and involvement of experts. In addition, those with IQs or DQs of 50 to 69, 70 to 84, 85 to 114, and 114 or higher were classified as having an intellectual disability, borderline intelligence, standard intelligence, and high intelligence, respectively. Participants were also asked to answer five questions on the developmental coordination disorder section of the Preschoolers’ Check List of obscure disAbilitieS (KITA, 2018), a questionnaire used to screen for stuttering, tic disorder, specific learning disabilities, and developmental coordination disorder. Furthermore, they were asked to answer six questions on the Japanese version of the Kessler Psychological Distress Scale, which measures the mental distress of respondents (Furukawa et al., 2008).

### Procedure

Participants were recruited through the mailing lists of “Hattatsu Navi,” a website providing information on developmental disorders with over 300,000 members; “Litalico Junior,” a child development support facility with 147 offices in Japan; and “Litalico Works,” a work support facility with 137 offices in Japan (Inc., 2023, 2024). The URL of the online platform (Questant) was sent to the participants who applied by email, and they were asked to respond. With the permission of the ethics review committee, informed consent was obtained through the platform. The sample size for this study was planned considering the recommendations of previous studies (Nylund-Gibson et al., 2023), which emphasize the importance of robust sample sizes for latent transition analysis to ensure stable estimation and accuracy. Based on this guidance, we aimed to secure over 300 participants to align with the data requirements of the simultaneous study by (Nakamura, 2024). Additionally, the sample size meets the COSMIN standard, which suggests that a sample size of at least 100 participants is sufficient for evaluating the reliability and responsiveness of measurement properties (Prinsen et al., 2018).

To maintain data quality, we implemented exclusion criteria for cases where participants selected the same response option for all 36 items on the PQP questionnaire. This approach was adopted to identify potential careless or non-differentiated responses, which could reduce data validity. Such cases were reviewed and subsequently excluded from the analysis, ensuring that only meaningful and valid data were included in the study.

The scales answered at each point by the participants are shown in Figure 1. The first data collection was conducted from July to February at four locations to avoid the institutional transition period in April when the Japanese school year begins. The first data collection was conducted from July to August 2024, and the URL for the online platform was sent to the email addresses of the research participants. The second data collection was conducted one week after the first for each participant, and the URL for the online platform was sent. The third data collection occurred three months after the first (October to November), and the fourth data collection occurred three months after the third (January to February). In consideration of the possibility that some participants may have been registered on multiple mailing lists, we informed them at the time of informed consent that they could not participate in the study more than once. After the data collection was completed, the first and second authors confirmed that there were no duplicate data based on the information regarding the child’s date of birth, prefecture of residence, and date of diagnosis of ASD.

**Figure 1.**
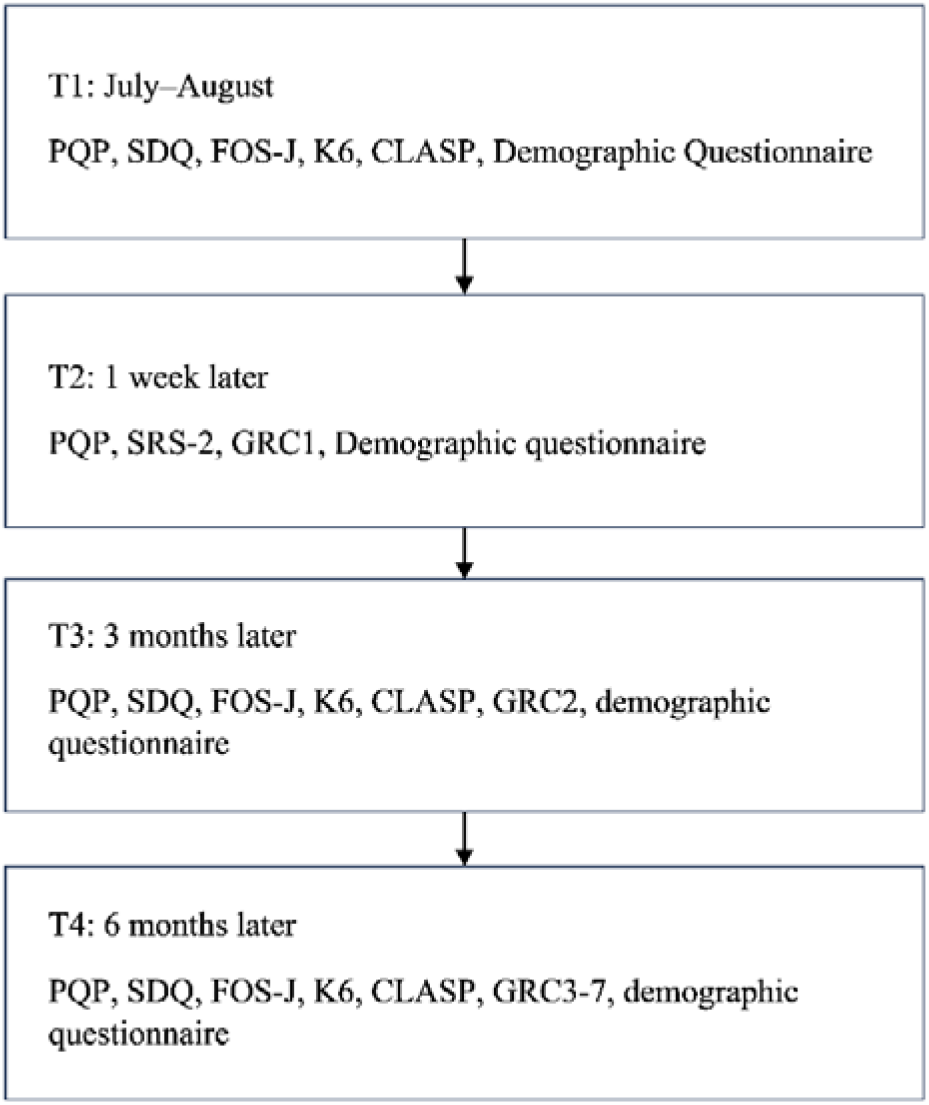
Data collection procedure. *Note.* PQP, Parental Questionnaire for Participation; SDQ, Strengths and Difficulties Questionnaire; FOS-J, Japanese version of the Family Outcomes Survey-Revised; K6, Kessler Psychological Distress Scale; CLASP, Checklist for obscure disabilities in Preschoolers; GRC, Global Rating of Change; SRS-2, Social Responsiveness Scale, Second Edition.

### Statistical Analysis

All statistical analyses were conducted on participants with complete data for the PQP, SDQ, FOS-J “Family Outcomes,” and demographic questionnaires at all time points from T1 to T4 (Figure 2). Therefore, there were no missing values in the analysis of this study.

**Figure 2.**
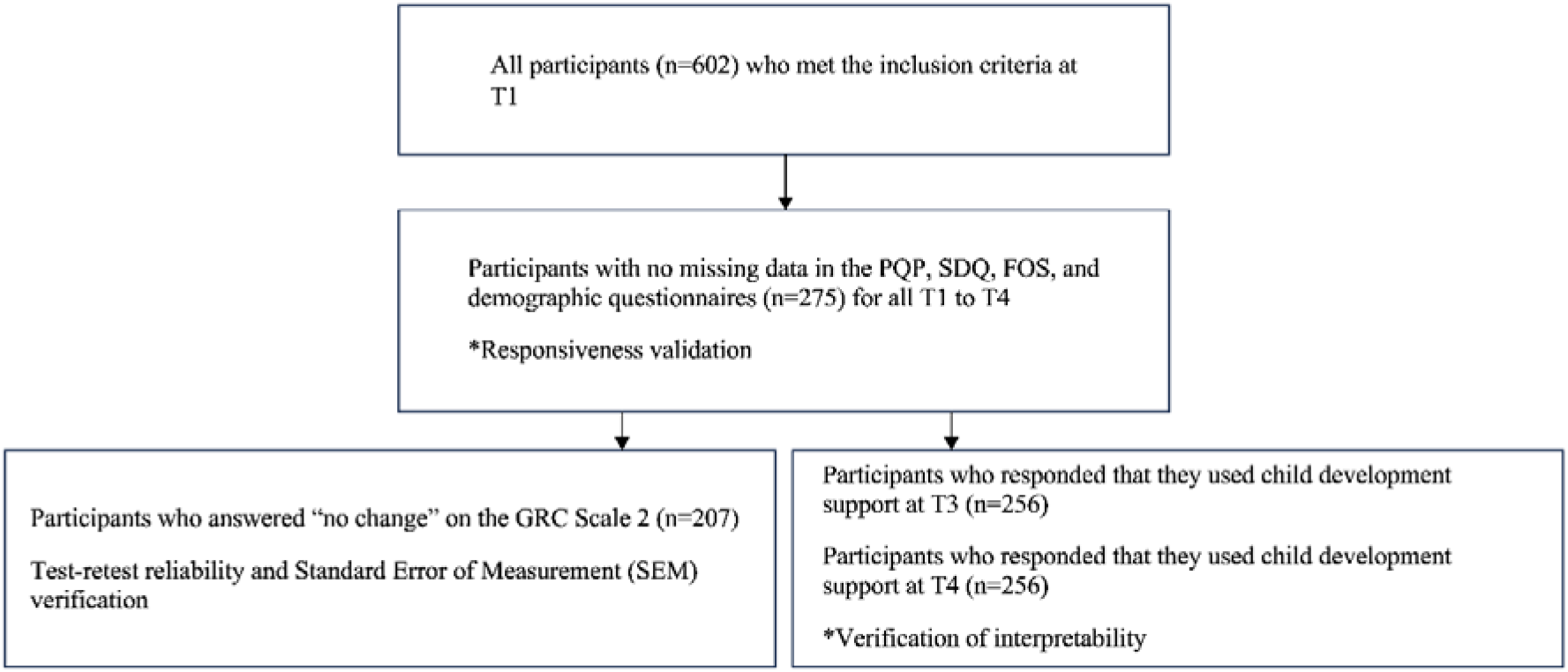
Participants of each statistical analysis.

#### Test−retest Reliability and Measurement Error

Test−retest reliability was assessed by calculating the intraclass correlation coefficient (ICC) for the total PQP score and each factor score obtained at T1 and T2 for participants who responded “4: No change” on the GRC Scale1 (Figure 2). An ICC of 0.7 or higher indicated sufficient reliability (Terwee et al., 2021). In addition, we used the Bland-Altman method to examine the limits of agreement (LOA) between the two measurement results. The LOA was calculated as “the mean difference between the two measurement results ± 1.96 × the standard deviation of the difference.” In this study, a Bland-Altman plot was created, with the difference between the two measurement results on the Y-axis and the average of the two measurement results on the X-axis. In this plot, if the difference between the two measurement results falls within this range, it is interpreted that there is consistency and stability between the two measurement results, as 95% of the data will fall within this range if the error follows a normal distribution.

The Standard Error of Measurement (SEM) was calculated according to the ICC as “SEM = SD × √(1−ICC)”. Further, the Minimal Detectable Change (MDC), which indicates the minimum change that can be detected as clinically significant, was calculated according to the SEM value, considering the effect of measurement error. The MDC was calculated as “MDC = 1.96 × SEM × √2.”

#### Responsiveness

Responsiveness was assessed for all participants (Figure 2). Specifically, reactivity was assessed through the following four hypothesis tests for changes in scores. These hypothesis tests were conducted twice for the change from T1 to T3 and the change from T3 to T4.

*Hypothesis 1: A negative correlation exists between the change in the total difficulty score of the SDQ and the change in the total score of the PQP*.

Problematic behavior in children with ASD is a barrier to participation in community life (Howell & Pierson, 2010), and multiple subdomains of the SDQ are negatively associated with participation in out-of-school activities in children with physical disabilities and typically developing children (King et al., 2013). Therefore, we assume that changes in the total difficulty score of the SDQ will show a weak to moderate negative correlation with changes in participation; further, the total PQP score and the total difficulty score of the SDQ in this study will show a correlation in the range of 0.2–0.5.

*Hypothesis 2: A negative correlation exists between the change in the SDQ peer relationship score and the change in the PQP total score*.

Problematic behavior in friendships is a construct that is expected to negatively correlate with the PQP Factor 1 (Nakamura, Nagayama, et al., 2024). However, since changes in problematic behavior in friendships do not necessarily correlate with changes in other factors, they should show a weak correlation with changes in the total PQP score. Therefore, we assume that they will show a negative correlation in the range of 0.1–0.3.

*Hypothesis 3: A positive correlation exists between the change in the Prosocial Behavior score on the SDQ and the change in the total PQP score*.

Prosocial Behavior is a concept that is related to core symptoms of ASD (Russell et al., 2013). Participation by children with ASD is related to core symptoms, and it is expected that the acquisition of Prosocial Behavior is partially related to milder core symptoms. Therefore, the change in Prosocial Behavior should show a weak to moderate positive correlation with the change in the total PQP score. Therefore, we assume that it will show a positive correlation in the range of 0.2–0.5.

*Hypothesis 4: A positive correlation exists between the total FOS-J score and the total PQP score and between the change in these scores*.

According to previous research, family functioning is predicted to positively correlate with the leisure and community participation of children with disabilities (King et al., 2013; Nakamura, Nagayama, et al., 2024). However, several results also show no correlation with friendships, family life, or participation in education (Di Marino et al., 2018; Nakamura, Nagayama, et al., 2024). Although the FOS-J is not a scale for measuring family functioning, it is a scale that can measure the contribution of early intervention in families, and it measures a similar construct (Bailey et al., 2008). Therefore, it is expected that the range of correlation for change would be slightly wider and there will be a relatively strong correlation with Factor 4, which includes content related to leisure and leisure activities, with a range of 0.0–0.3 for the change in the total score of PQP.

All correlations were calculated using Spearman’s rank correlation coefficient.

#### Interpretability

We calculated the MIC, which indicates the threshold at which a measurement tool can detect the minimum clinically meaningful change (Prinsen et al., 2018). The MIC is an essential criterion for determining the extent to which the effect of an intervention is clinically significant, but it depends on the characteristics of the target population and intervention, and the estimated value often has a range that differs depending on these backgrounds. Therefore, for the MIC to function properly, it is necessary to calculate and verify it based on the context of the intervention for the target population that has received the specific intervention (Revicki et al., 2008). Therefore, we verified the MIC for children who use child development support (Ministry of Health, Labour and Welfare, 2020), a welfare service widely and publicly provided to preschool children with neurodevelopmental disorders in Japan (Figure 2).

The MIC was estimated using the receiver operating characteristic (ROC)-based method (MICROC), one of the anchor-based methods recommended by Terwee et al. (2021). The GRC Scale 2 to 7 was used as the anchor. As the first step in estimating the MIC value, the correlation coefficient between the change in the PQP and the anchor was calculated using Spearman’s rank correlation coefficient. This first step was conducted to verify the validity of the anchor, and the criterion value for the correlation coefficient was set at 0.30 or higher (Devji et al., 2020). Next, to estimate the MIC value for PQP improvement, an analysis using the ROC method was conducted on participants with an anchor GRC score of 4 (no change) or higher. In the ROC method, the Youden index was calculated according to the sensitivity and specificity of PQP as follows: “Youden index = sensitivity + specificity - 1 (Youden, 1950).” In this study, we considered that the highest Youden index represented the optimal PQP change score that optimally distinguished between the optimal MIC values, that is, 5 (slightly improved) and 4 (no change). The area under the curve (AUC) of the ROC curve was evaluated as excellent if the AUC value was ≥0.90, good if it was 0.80–0.89, acceptable if it was 0.70–0.79, and poor if it was less than 0.70 (Metz, 1978).

IBM SPSS Statistics Version 25 was used for all statistical analyses.

## Results

A total of 545 participants who met the inclusion criteria at Time Point 1 participated in the study, and 275 (50.5%) completed the survey without dropping out until Time Point 4. Table 2 shows the demographic characteristics of participants who completed the survey until Time Point 4, and Table 3 shows the PQP, SDQ, and FOS-J scores at each time point.

**Table 2.**
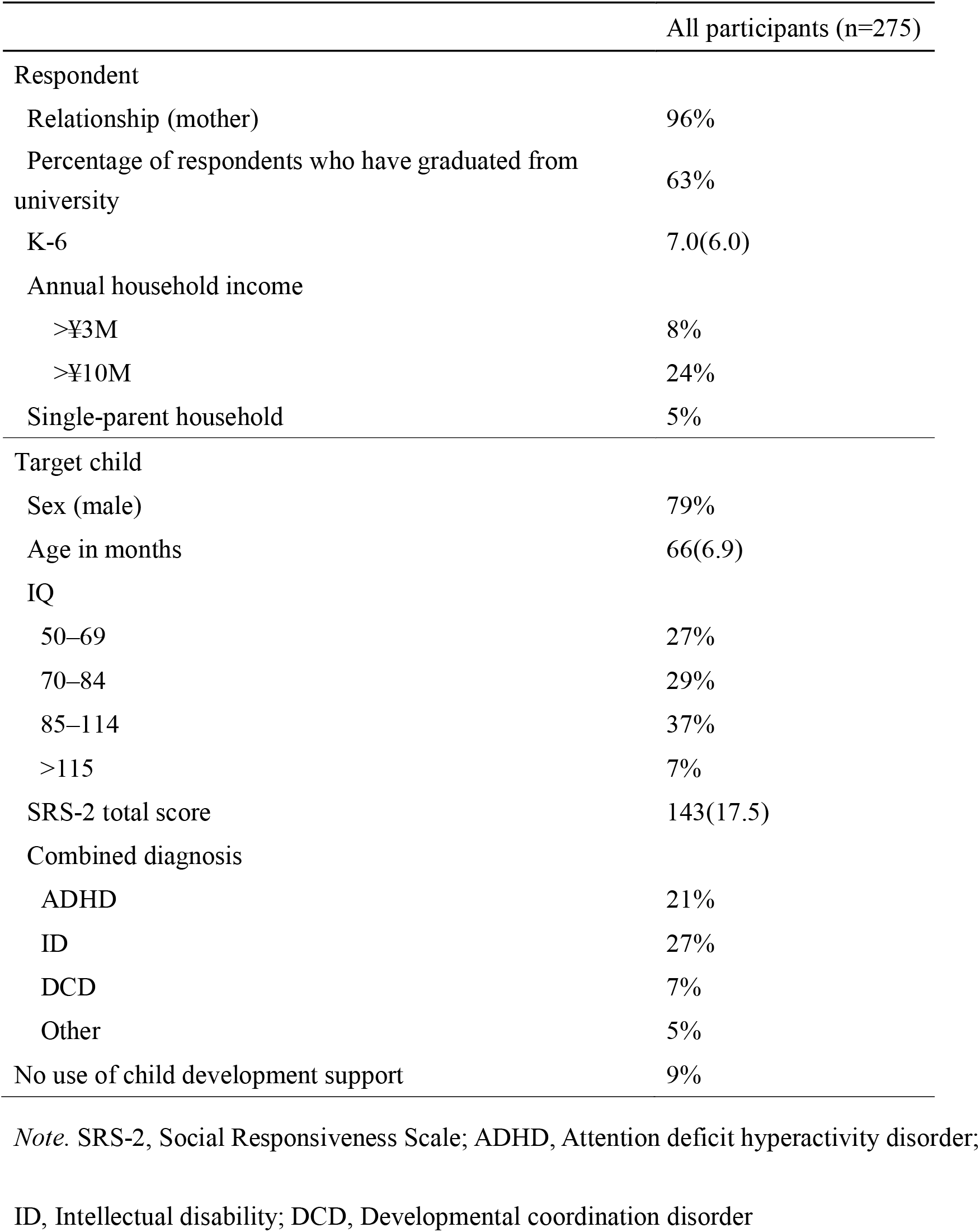
Participant Characteristics.

|  | All participants (n=275) |
| --- | --- |
| Respondent |  |
| Relationship (mother) | 96% |
| Percentage of respondents who have graduated from university | 63% |
| K-6 | 7.0(6.0) |
| Annual household income |  |
| >¥3M | 8% |
| >¥10M | 24% |
| Single-parent household | 5% |
| Target child |  |
| Sex (male) | 79% |
| Age in months | 66(6.9) |
| IQ |  |
| 50–69 | 27% |
| 70–84 | 29% |
| 85–114 | 37% |
| >115 | 7% |
| SRS-2 total score | 143(17.5) |
| Combined diagnosis |  |
| ADHD | 21% |
| ID | 27% |
| DCD | 7% |
| Other | 5% |
| No use of child development support | 9% |

*Note.* SRS-2, Social Responsiveness Scale; ADHD, Attention deficit hyperactivity disorder;
ID, Intellectual disability; DCD, Developmental coordination disorder

**Table 3.** Scores at Each Time Point for the PQP, SDQ, and FOS-J.

|  | T1 | T2 | T3 | T4 |
| --- | --- | --- | --- | --- |
| factor1 | 24(7.5) | 24(6.8) | 24(6.6) | 25(7.0) |
| factor2 | 26(5.3) | 25(5.3) | 26(5.3) | 26(5.2) |
| factor3 | 25(5.5) | 25(5.3) | 26(5.1) | 25(5.4) |
| factor4 | 31(5.5) | 32(5.6) | 32(5.4) | 32(5.6) |
| PQPtotal | 104(18.2) | 104(17.6) | 106(16.8) | 108(17.5) |
| SDQ-PP | 9(2.2) | - | 9(2.2) | 8(2.2) |
| SDQ-PB | 4(2.6) | - | 4(2.5) | 4(2.4) |
| SDQ-TDS | 32(5.3) | - | 31(5.1) | 31(5.3) |
| FOS-J total | 74(17.8) |  | 74(16.7) | 73(17.1) |

*Note.* PQP, Parental Questionnaire for Participation; SDQ, Strengths and Difficulties
Questionnaire; FOS-J, Japanese version of the Family Outcomes Survey-Revised.

### Test−retest Reliability and Measurement Error

Of the respondents, 207 (75% of the total) answered that there had been no change in their daily lives according to the GRC Scale 1. Table 4 shows the ICC, means, SEM, and 95% confidence intervals for the PQP subscale and total scores for these 207 respondents at T1 and T2. Figure 3 shows the results of the Bland-Altman analysis. The ICC for the total PQP score was 0.93, which exceeded 0.7. The ICC for each factor score of the PQP ranged from 0.80 to 0.92, and all exceeded 0.7. Further, the Bland-Altman method showed that most of the measurement results for the total score and each factor score fell within the LOA. The SEM, calculated on the basis of the ICC, was 4.78 for the total score and ranged from 2.09 to 2.42 for each factor score. Additionally, the MDC, calculated on the basis of the SEM, was 13.25 for the total score, with each factor score ranging from 5.79 to 6.71.

**Figure 3.**
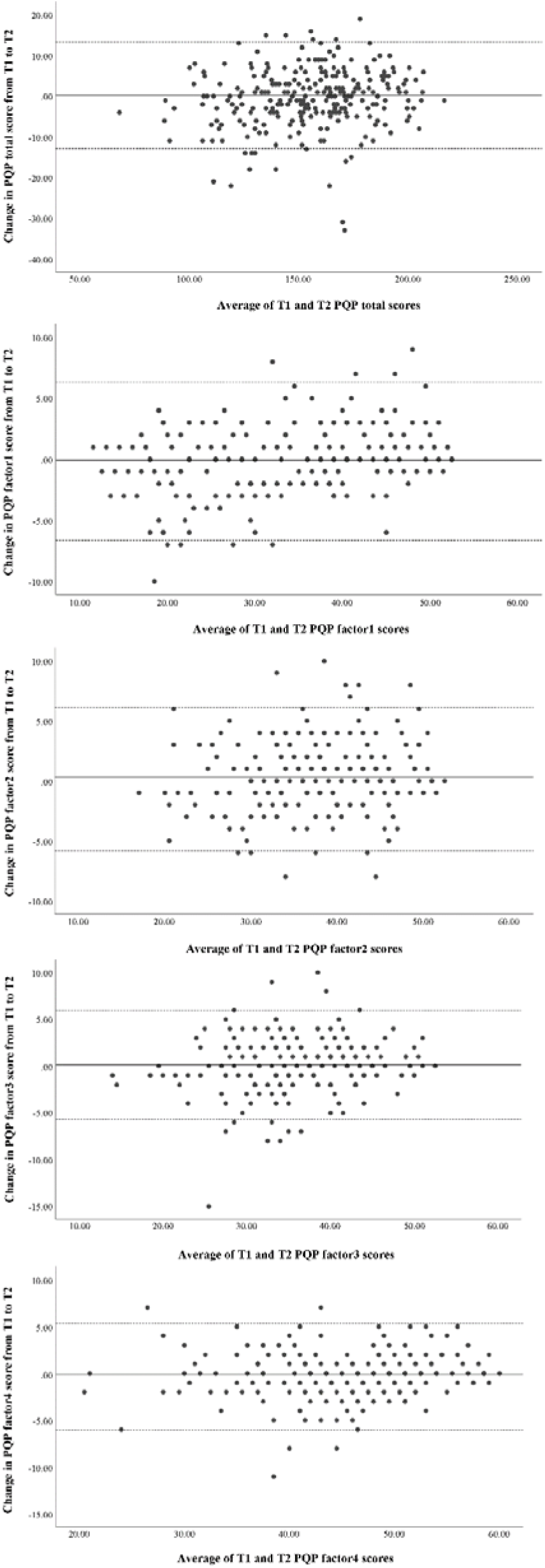
a. Bland-Altman plot for the PQP total score. b. Bland-Altman plot for the PQP Factor 1 score c. Bland-Altman plot for the PQP Factor 2 score d. Bland-Altman plot for the PQP Factor 3 score e. Bland-Altman plot for the PQP Factor 4 score.

**Table 4.**
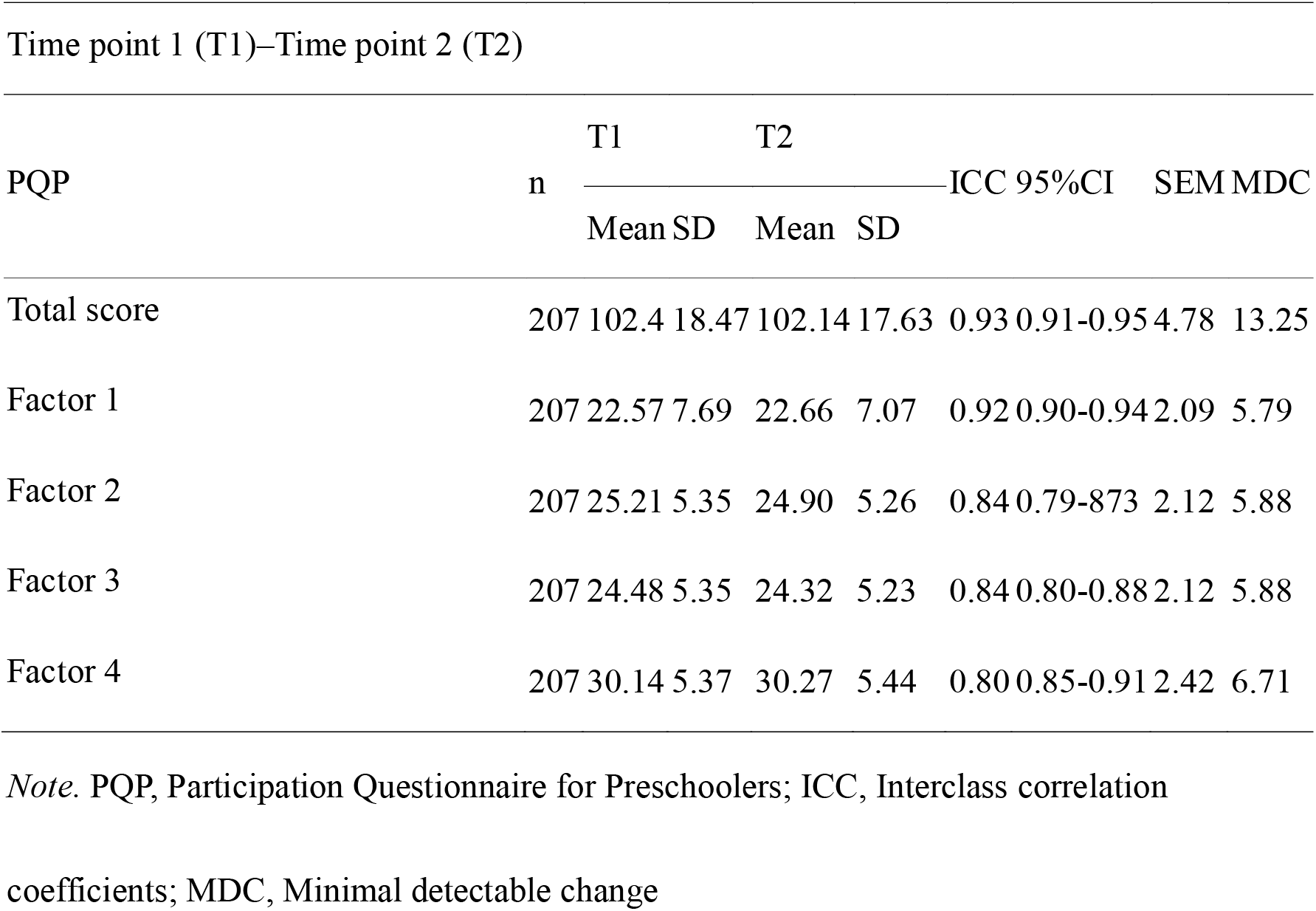
Test−Retest Reliability and Standard Error of Measurement Verification Results.

### Responsiveness

Table 5 shows the correlation coefficients for the changes in the total score and each factor of the PQP for all 275 respondents, as well as the correlation coefficients for the changes in age, SDQ-TDS score, SDQ-PP score, and FOS-J total score from T1 to T3 and T3 to T4. Only the results regarding the correlation between the change in prosociality on the SDQ and the PQP (Hypothesis 3) were outside the expected range. A post-hoc analysis was conducted to investigate the reason for this unexpected result. The correlation between the Prosocial Behavior score and the SRS-2 total score, which underpinned our hypothesis, was calculated using Spearman’s rank correlation coefficient, yielding a value of −0.223.

**Table 5.** Results of Hypothesis Testing for Responsiveness Validation.

| T1–T3 | PQP<br>total | PQP<br>Factor1 | PQP<br>Factor2 | PQP<br>Factor3 | PQP<br>Factor4 |
| --- | --- | --- | --- | --- | --- |
| SDQ-TDS | -0.288** | -0.250** | -0.108 | -0.136* | -0.052 |
| SDQ-PP | -0.275** | -0.348** | -0.095 | -0.155* | 0.006 |
| SDQ-PB | 0.112 | 0.143* | 0.018 | 0.120* | 0.081 |
| FOS-J | 0.046 | 0.014 | 0.013 | 0.049 | 0.183** |

| T3–T4 | PQP<br>total | PQP<br>Factor1 | PQP<br>Factor2 | PQP<br>Factor3 | PQP<br>Factor4 |
| --- | --- | --- | --- | --- | --- |
| SDQ-TDS | -0.342** | -0.266** | -0.209** | -0.165** | -0.116 |
| SDQ-PP | -0.259** | -0.271** | -0.065 | -0.215** | -0.039 |
| SDQ-PB | 0.179** | 0.124* | 0.029 | 0.162** | 0.148* |
| FOS-J | 0.134* | 0.054 | 0.055 | 0.031 | 0.161** |

*Note.* \*\*p < 0.01, \*p < 0.05; PQP, Participation Questionnaire for Preschoolers; SDQ,
Strengths and Difficulties Questionnaire; FOS-J, Japanese version of the Family Outcomes
Survey-Revised.

### Interpretability

At T3 and T4, 256 respondents (93% of the total) reported that they were using child development support services. As the first step in calculating the MIC, we calculated the correlation coefficients between the GRC scales 2–7 and the total PQP score, as well as the scores for each factor. However, since all of these correlations fell below the predetermined threshold of 0.3, the calculation of the MIC using the MICROC method was not performed (Table 6). To investigate the factors contributing to the low correlations, we conducted a post-hoc analysis of the correlations between each GRC Scale and the changes in the scores of the scales used to assess responsiveness. The results showed that most of the correlation coefficients were close to zero compared with the PQP.

**Table 6.** Correlation Coefficients between Changes in Each Score and Changes in the GRC Scale.

| T1-T3 | GRC2 |
| --- | --- |
| PQP-Total | 0.195** |
| SDQ-TDS | -.089 |
| SDQ-PP | -.003 |
| SDQ-PB | -.003 |
| FOS-J | .012 |

| T1-T3 | GRC3 | GRC4 | GRC5 | GRC6 | GRC7 |
| --- | --- | --- | --- | --- | --- |
| PQP-Total | .229** |  |  |  |  |
| PQP-Factor1 |  | .175** |  |  |  |
| PQP-Factor2 |  |  | .056 |  |  |
| PQP-Factor3 |  |  |  | .048 |  |
| PQP-Factor4 |  |  |  |  | .107 |
| SDQ-TDS | -.235** | -.244** | -.218** | -.160* | -.221** |
| SDQ-PP | -.123* | -.151* | -.045 | -.052 | -.075 |
| SDQ-PB | .047 | .096 | .016 | -.017 | .062 |
| FOS-J | .071 | .021 | .058 | .011 | .039 |

*Note.* \*\*p < 0.01, \*p < 0.05. PQP, Participation Questionnaire for Preschoolers; SDQ,
Strengths and Difficulties Questionnaire; FOS-J, Japanese version of the Family Outcomes
Survey-Revised; GRC, Global Rating of Change.

## Discussion

### Reliability

The ICC results and measurement error values suggest that the PQP has high reproducibility and that no bias exists toward positive or negative values in the Bland-Altman plot. This indicates that the effects of fixed and proportional errors are small and, despite random errors, consistency is high. These results confirm that the PQP demonstrates high reproducibility in the total and factor scores, with minimal variation in results. To the best of our knowledge, the longitudinal reliability of existing disorder-specific measurement tools for ASD has not been verified (Castro & Pinto, 2015; Golos et al., 2022; Li et al., 2023). Further, while the interval periods differ across tools, the ICC of the PQP was generally comparable to, or better than, that of other widely used participation measurement tools (Rosenberg et al., 2010); in some cases, it performed even better (Bult et al., 2013; Coster et al., 2011; Kang et al., 2017; Khetani et al., 2015; Nordtorp et al., 2013). Regarding reliability, the PQP offers advantages in measuring participation in children with ASD.

### Responsiveness

In this study, we tested four hypotheses twice to examine responsiveness, and the results aligned with the hypotheses in six of the eight tests. The direction, strength, and trends of the factor scores for the SDQ’s TDS and PP scores, as well as the FOS-J score, fell within the expected range. However, although the correlation for the SDQ’s PB score was in the expected direction, the strength of the correlation was lower than anticipated. This may be due to the hypothesis’s basis—a negative correlation between ASD core symptoms and prosociality—being weaker than expected. Nevertheless, with 75% of the initial hypotheses falling within the expected range, the PQP demonstrates generally good responsiveness in longitudinally measuring participation. While recent studies, such as Soref et al. (2023), have increasingly evaluated the effects of interventions on children’s participation, most major participation measurement tools still lack responsiveness validation (Bult et al., 2013; Coster et al., 2011; Kang et al., 2017; Khetani et al., 2015; Nordtorp et al., 2013). In this context, the PQP’s acceptable level of responsiveness represents a significant advantage for its application.

### Interpretability

We attempted to validate the MIC using anchors; however, the correlation coefficients necessary to calculate the MIC for all six anchors did not meet the required threshold. This could be due to several factors. Regarding reliability, since the measurement error of the PQP was good, it is possible that the measurement error of the GRC Scale reduced the correlation coefficient. Regarding content validity, the construct of participation is complex and multidimensional; therefore, it is possible that a single-item question was insufficient to capture adequate correlations.

### Limitations

This study had three limitations. First, over 50% of participants dropped out before completing the study, and those who completed the responses had a higher rate of university graduation compared with the average Japanese population (Ministry of Health, Labour and Welfare, 2022; Statistics Bureau, 2021). Therefore, the generalizability of the results may be limited. Second, the study targeted preschool children aged 51–75 months, excluding children aged 36–50 months. Consequently, the findings may not be generalizable to all preschool children aged 3–6 years, which is the full target age range of the PQP. Third, a prototype version of the PQP was used. Although items excluded from the final version were not included in the scoring, participants were still asked to respond to these items. The use of the prototype version may have impacted the evaluation of responsiveness and interpretability. Therefore, future research should validate the final version of the PQP by including younger age groups and caregivers from more diverse backgrounds.

### Conclusion

Although the MIC could not be calculated, the PQP had good reliability, low measurement error, and acceptable responsiveness. This suggests that the PQP is a useful measurement tool for practice, longitudinal observational studies, and intervention studies. However, to further enhance its applicability, future research should validate the final version of the PQP by including younger age groups and caregivers from more diverse backgrounds. Additionally, exploring its responsiveness across various intervention types and settings could provide a broader foundation for its use in diverse clinical and research contexts.

## Data Availability

The data that support the findings of this study are not publicly available due to restrictions imposed by the Kanagawa University of Human Services Research Ethics Committee. Data cannot be shared under any circumstances.

## Notes

### Competing Interest Statement

The authors have declared no competing interest.

### Funding Statement

This work was supported by the Japan Society for the Promotion of Science (JSPS) KAKENHI for the category of Basic Research C (Grant number 22K02410), awarded to Takuto Nakamura.

### Author Declarations

The Research Ethics Committee of Kanagawa University of Human Services approved this study (Approval Number: 31-14-009). Participants were provided with the URL of the online platform (Questant) via email, and informed consent was obtained through the platform.

